# What factors converged to create a COVID-19 hot-spot? Lessons from the South Asian community in Ontario

**DOI:** 10.1101/2022.04.01.22273252

**Authors:** Sonia S Anand, Corey Arnold, Shrikant Bangdiwala, Shelly Bolotin, Dawn Bowdish, Rahul Chanchlani, Russell de Souza, Dipika Desai, Sujane Kandasamy, Farah Khan, Zainab Khan, Marc-André Langlois, Jayneel Limbachia, Scott Lear, Mark Loeb, Lawrence Loh, Baanu Manoharan, Kiran Nakka, Martin Pelchat, Zubin Punthakee, Karleen Schulze, Natalie Williams, Gita Wahi

**Author notes:** **Address for Correspondence:** Dr. Sonia Anand, Chanchlani Research Centre, Population Health Research Institute, 1280 Main St West, McMaster University. **Declaration of competing interests:** M. L. has received vaccine advisory board consulting fees for Seqirus, Pfizer, Merck, Sanofi, Medicago; grant funding for vaccine trial from Seqirus; in-kind vaccine from Sanofi for a trial; and is on the Vaccine Data Safety Monitoring board for Medicago, NIH, CanSino biologics.

## Abstract

**Background:** South Asians represent the largest non-white ethnic group in Canada. The Greater Toronto Area (GTA), home to a high proportion of South Asians, emerged as a COVID-19 hot spot. Early in the pandemic, the South Asian community was identified as having risk factors for exposure and specific barriers to accessing testing and reliable health information, rendering them uniquely vulnerable to SARS-CoV-2 infection.

**Objectives:** To investigate the burden of SARS-CoV-2 infection among South Asians in the GTA, and to determine which demographic characteristics were most closely aligned with seropositivity, in this cross-sectional analysis of a prospective cohort study.

**Methods:** Participants from the GTA were enrolled between April and July 2021. Seropositivity for anti-spike and anti-nucleocapsid antibodies was determined from dried blood spots, and age and sex standardized to the Ontario South Asian population. Demographics, risk perceptions, and sources of COVID-19 information were collected via questionnaire in a subset.

**Results:** Among the 916 South Asians enrolled, mean age 41 years, the age and sex standardized seropositivity was 23.6% (95% CI: 20.8%-26.4%). Approximately one-third identified as essential workers, and 19% reported living in a multi-generational household. Over half perceived high COVID-19 risk due to their geographic location, and 36% due to their type of employment. The top three most trusted sources of COVID-related information included healthcare providers/public health, traditional media sources, and social media.

**Conclusion:** By the third wave of the COVID-19 pandemic, approximately one-quarter of a sample of South Asians in Ontario had serologic evidence of prior SARS-CoV-2 infection. Insight into factors that render certain populations at risk can help future pandemic planning and disease control efforts.

## Background

The novel SARS-CoV-2 virus was declared a global pandemic in March 2020. Within Canada, COVID-19 hotspots emerged, and attention was drawn to these regions by the high infection and hospitalization rates and need to transfer patients outside of these regions to receive intensive care. South Asians (i.e., people who originate from the Indian subcontinent) are the largest non-white ethnic group in Canada (1) and have been disproportionally affected by COVID-19 (2). More than half of the residents of the Region of Peel, in the Greater Toronto Area (GTA), Ontario identify as South Asian (3).

Peel is divided into three cites - Caledon, Brampton, and Mississauga. Brampton has a population of 600,000; and over one-third of residents are South Asian, rising in some areas to nearly two thirds (4). In the first wave of the COVID-10 pandemic in Ontario [March to September 2020], the SARS-CoV-2 infection rate was almost twice as high in the City of Brampton, compared to the rate in neighbouring Mississauga (711 vs 390 cases per 100,000 people) (2). Peel emerged as a hotspot in Ontario, accounting for 22% of provincial cases during the second wave, beginning in September 2020 (2,5), although making up only 10% of the province’s population (Figure 1), with the City of Brampton as the epicentre. Public Health Ontario provided indirect measures of the impact on the South Asian populations, showing the rates of infection, hospitalization, intensive care unit admission and death rates by quintiles of diversity in Ontario. People in the most diverse neighbourhoods were more likely to be new immigrants, younger, living in larger households, and had three times higher infection, four times the hospitalization, four times the intensive care unit admission, and two times the death rate from COVID-19 as compared to less diverse communities (6). Early in the pandemic, there was limited availability of SARS-CoV-2 laboratory testing throughout Ontario. In hotspot communities, like Peel, this was compounded by other access barriers to health care which pre-dated the pandemic, which resulted in a high burden of infection (2). However, action by local and provincial public health officials, combined with advocacy from community groups and the media (7) resulted in increased resources for testing and prioritization of vaccine roll-out, which commenced in April 2021 (8).

**Figure 1:**
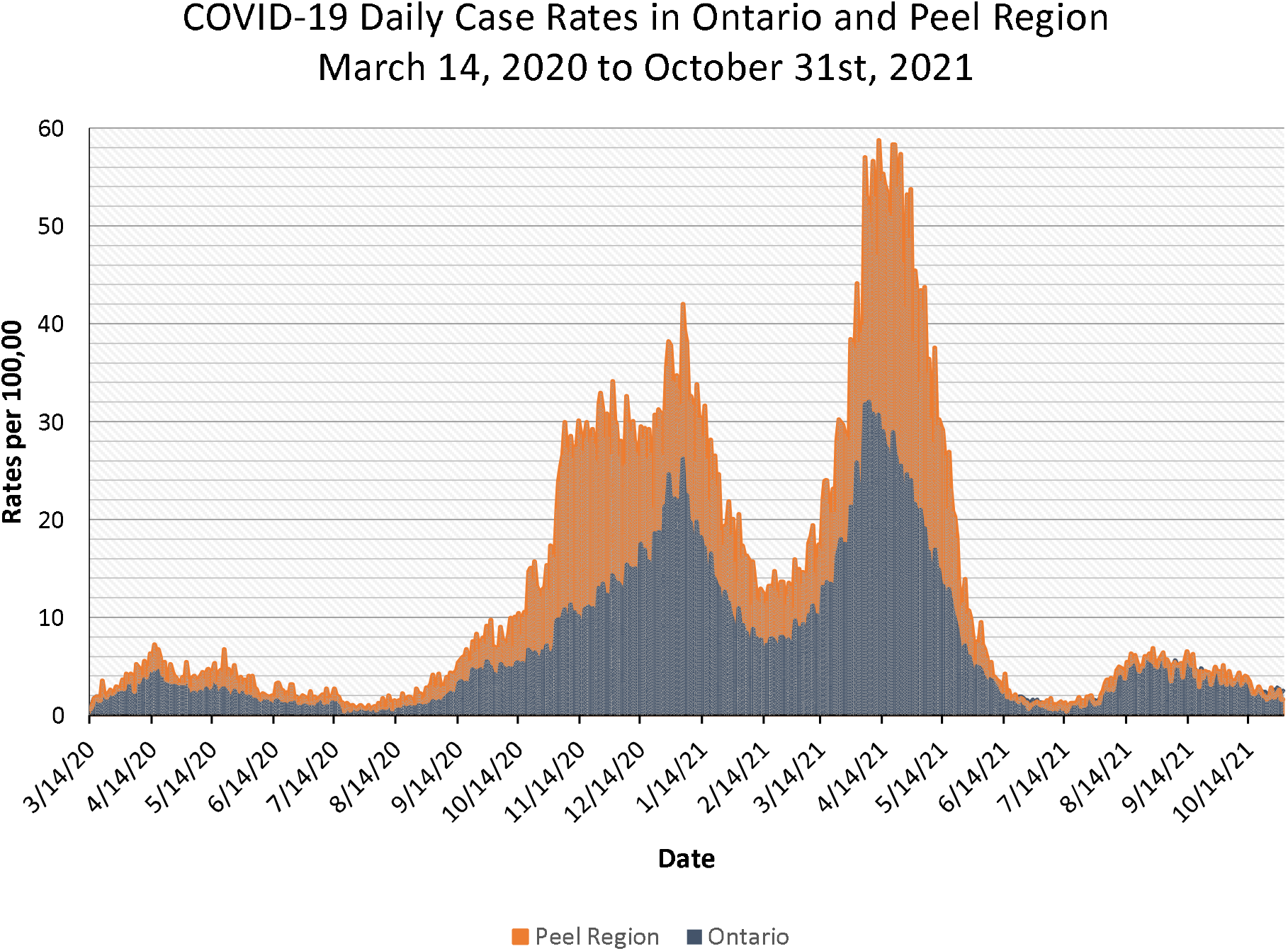
Data from Public Health Ontario: COVID-19 Pandemic Comparing Peel Region to Ontario (30).

Here we report the seroprevalence of SARS-CoV-2 infection in this ethnic group who were recruited during Wave 3 of the pandemic, [March 2021 to June 2021] and report the factors which correlate with infection.

### Design and Methods

The COVID CommUNITY study is a prospective cohort study focused on South Asian adults living in Canada, in the provinces of Ontario and British Columbia. In this cross-sectional analysis, we present data from the baseline assessment of the Ontario sub-cohort recruited between April 14, to July 28, 2021. All participants provided informed consent and this study was approved by Hamilton Integrated Research Ethics Board (13323 - March 24, 2021).

This study was funded by the COVID-19 Immune Task Force (CITF), and funds distributed from the Public Health Agency of Canada.

#### Study Population

Adults ≥ 18 years of age and South Asian were eligible. South Asian ethnicity was self-reported and defined by parental South Asian ancestry from the Indian subcontinent, Africa, Caribbean, and Guyana.

#### Recruitment

Recruitment was predominantly from vaccination centres in Brampton immediately after receiving dose 1 (Pre-vaccination group), or > 24 hours after receiving dose one or two of vaccination (Post vaccination group). A smaller proportion came from places of worship, SARS-CoV-2 testing centre in Brampton, through social media, and by inviting participants of South Asian origin from an existing cohort study (9). At the time of consent, dried blood spots (DBS) were collected along with key sociodemographic information, and vaccination status. Additional information regarding employment type, health history, prior SARS-CoV-2 infection, perception of SARS-CoV-2 infection risk, primary and trusted sources of COVID-19 information was collected on a web-based survey.

#### Laboratory measurements

DBS were collected in person at recruitment sites, and through the mail for social media and cohort recruitment. For home collection, an instructional video and self-addressed stamped envelope was provided for mail returns. Blood was collected on the WhatmanTM 903® Protein Saver Cards. Batched specimens were sent for analysis to the CITF-funded lab of our collaborator (Dr. Langlois) for analysis and were processed and analyzed using a high throughput ELISA assay (10,11). The main assays focused on the parallel detection of immunoglobulin IgG against the spike trimer (S), and the nucleocapsid (N) protein with standardized antigens to S, and N produced by the National Research Council of Canada laboratory. This method has been used in prior studies (12), and detailed methods are described elsewhere (12,13). The IgG-based ELISAs measure antibody levels in single-point measurements in reference to a standard antibody curve to accurately distinguish individuals with previous infection or vaccination from those who have not been infected or vaccinated (Area Under the Receiver Operating Curve > 0.96 for each assay).

Evidence of previous infection was defined in pre-vaccinated participants (i.e. – those unvaccinated or those submitting samples immediately after receiving vaccine) as individuals with test results having signal to cutoff ratios ≥1 for both anti-S and anti-N IgG, or anti-S≥ 1 and a history of SARS-CoV-2 infection. Evidence of previous infection among vaccinated individuals (i.e. those who submitted a DBS sample >1 day after vaccination) was defined as an anti-N IgG results of ≥ 1. A range of strict definition and lenient definition in the pre-vaccination group was also examined and defined as a) IgG S <u>and</u> IgG N ≥ 1, and b) IgG S ≥ 1, respectively. The false discovery rate (FDR) was set at 2% for the IgG S, and 3% for IgG to N; These cut-offs have > 98% accuracy when validated by the National Microbiology Laboratory reference panel (11). The background seasonal coronavirus antibody mean titers in the Ottawa region was used as reference (10,13).

### Statistical Analysis

The proportion of participants who had evidence of prior SARS-CoV-2 infection is compared to the infection rate in similar forward sortation areas (FSA). Risk factors for SARS-CoV-2 infection, sources of health information, and perception of SARS-CoV-2 risk are reported as percentages or means (SD) for the overall cohort, and for comparing seropositive to non-infected participants. Non-responders to the follow-up survey were compared to responders for age, sex, income, location, and seroprevalence. As this sample was not a random sample of the population, the risk of bias was assessed with a modified Joanna Briggs instrument (JBI) Critical Appraisal Checklist for Prevalence Studies (14). This tool evaluates the external and internal validity of the study using the CocoPop mnemonic (Condition, Context, Population) (15). The overall risk of bias was determined based on the criteria met and the impact any criteria that were not met have on the validity and reliability of the prevalence estimate (14). Items include questions on the sampling frame, sampling method, sample size, coverage, validity, and reliability of the condition measured, statistical analysis, and response rate, as shown in Supplementary Table 1.

## Results

Recruitment into the COVID CommUNITY study in Ontario began on April 14, 2021, and 939 participants recruited, and a DBS collected up to July 28, 2021. Of 939 participants, 916 participants DBS were analyzed, and 75.7% of these participants completed the follow-up web-survey (Supplementary Figure 1). Participants who did not complete the survey were older, and more likely to be seropositive, although no significant differences in location of recruitment, household income, or sex were observed (Supplementary Table 2).

### Demographic Characteristics

Characteristics of the overall cohort are shown in Table 1. The average age of participants was 41.5 years (14.4), and 49% were women. Briefly, over 90% of participants lived in the Peel Region, the majority (82%) from the City of Brampton. About two-thirds of participants were born in Canada or had lived in Canada more than 10 years. Most participants (78%) had completed post-secondary education, and 72% were employed. Almost 33% of participants’ jobs were classified as essential work using the Ontario Government classification (i.e., food manufacturing and transportation workers), and an additional 16% of participants preferred not to answer this question. Although multigenerational household data were incomplete, of the 576 participants who completed this section, 19% reported living in a multigenerational household, with an additional 12% who preferred not to answer. The most common mother tongue reported included Punjabi or Urdu, followed by Gujarati, and Hindi. Demographic characteristics by pre- or post vaccination status are found in Supplementary Table 3.

**Table 1:** Demographic Characteristics.

|  | Percentage % | Responses |
| --- | --- | --- |
| **Overall** |  | 916 |
| **Sex** |  | 916 |
| Female | 49.2 |  |
| Male | 50.4 |  |
| Self-described | 0.3 |  |
| **Age group** |  | 906 |
| 18-24 | 10.5 |  |
| 25-34 | 25.7 |  |
| 35-44 | 27.3 |  |
| 45-54 | 17.3 |  |
| 55-64 | 10.4 |  |
| 65+ | 8.8 |  |
| **Vaccinated** |  | 916 |
| Yes, 2 doses | 7.1 |  |
| Yes, 1 dose | 42.9 |  |
| No | 50.0 |  |
| **Self Reported History of previous COVID-19 infection** |  | 699 |
| Yes | 12.6 |  |
| No | 85.8 |  |
| Unknown | 1.6 |  |
| **Median Household Income (2015) based on FSA** |  | 904 |
| \$40-<\$60K | 1.0 | |
| \$60-<\$80K | 10.7 | |
| \$80-<\$100K | 63.1 | |
| \$100K+ | 25.2 | |
| Prefer not to answer | 0.0 |  |
| **Essential workers #** |  | 693 |
| Yes | 32.9 |  |
| No | 50.8 |  |
| Prefer not to answer | 16.3 |  |
| **Completed Education** |  | 693 |
| High school or less | 17.5 |  |
| College, Trade, Certificate | 12.8 |  |
| University degree | 65.4 |  |
| Prefer not to answer | 4.3 |  |
| **Multi-generational Household** |  | 654 |
| Yes | 19.1 |  |
| No | 69.0 |  |
| Prefer not to answer | 11.9 |  |
| **Years in Canada** |  | 717 |
| 10 years or less | 31.1 |  |
| >10 years | 53.6 |  |
| Born in Canada | 11.9 |  |
| Prefer not to answer | 3.5 |  |
| **Mother tongue\*** |  | 732 |
| Punjabi or Urdu | 49.6 |  |
| Hindi | 11.3 |  |
| Gujarati | 14.5 |  |
| Other South Asian Languages | 20.8 |  |
| English | 7.4 |  |
| Prefer not to answer | 0.5 |  |
| **Medical history** |  |  |
| CVD (MI/Angioplasty/Stroke) | 3.1 | 679 |
| **Chronic Medical condition requiring medication** |  | 666 |
| Hypertension | 7.2 |  |
| Diabetes | 8.0 |  |
| Arthritis | 1.5 |  |
| Chronic Lung disease | 0.2 |  |
| Cancer | 0.2 |  |
| **Smoking Status** |  | 641 |
| Never | 86.6 |  |
| Former | 8.3 |  |
| Current | 5.1 |  |
| **Location** |  | 905 |
| Brampton Resident | 82.0 |  |
| Peel Resident | 91.7 |  |

\*Multiple answers can be selected. CVD: Cardiovascular Disease

### Seropositivity

The proportion of seropositive cases by geographic region (FSA) are shown in comparison to the cumulative incidence of SARS-CoV-2 infection rate (Figure 2). The age-sex standardized seropositivity for previous infection was 23.6% (95% CI: 20.8 to 26.4). Among the 458 participants without a prior vaccination, the seroprevalence was 26.9% (95% CI: 22.8 to 31.0) compared to 21.3% (95% CI: 17.5 to 25.1) in the 458 participants who had received at least one dose of the COVID-19 vaccine (393 received one, 65 had received 2 doses). The seropositivity ranged from 22.7% applying the strict definition of seropositivity, to 27.4% using the lenient definition. The risk of bias assessment was classified as moderate, attributed to the complex, multiprong, non-probability sampling method of recruitment (Supplementary Table 1). The seropositivity was higher in men, older ages, lower education, living in multigenerational households, and from the City of Brampton. Of participants who were seropositive, 53% did not report previous SARS-CoV-2 infection (Table 2).

**Table 2:** Seropositivity by Demographics.

|  | **Number of**<br>**samples**<br>**tested** | **Number of**<br>**positives** | **Seropositive**<br>**%** | **95% CI** |
| --- | --- | --- | --- | --- |
| **Overall** | 916 | 212 | 23.1 | 22.9 to 23.4 |
| **Sex** |  |  |  |  |
| Female | 451 | 96 | 21.3 | 20.9 to 21.7 |
| Male | 462 | 115 | 24.9 | 24.5 to 25.3 |
| Self-described | 3 | 1 | 33.3 | 28.0 to 38.7 |
| **Age group** |  |  |  |  |
| 18-24 | 95 | 22 | 23.2 | 22.3 to 24.0 |
| 25-34 | 233 | 49 | 21.0 | 20.5 to 21.6 |
| 35-44 | 247 | 54 | 21.9 | 21.3 to 22.4 |
| 45-54 | 157 | 36 | 22.9 | 22.3 to 23.6 |
| 55-64 | 94 | 28 | 29.8 | 28.9 to 30.7 |
| 65+ | 80 | 22 | 27.5 | 26.5 to 28.5 |
| **Vaccinated\*** |  |  |  |  |
| Yes | 458 | 107 | 23.4 | 23.0 to 23.7 |
| 2 doses | 65 | 12 | 18.5 | 17.5 to 19.4 |
| 1 dose | 393 | 93 | 23.7 | 23.2 to 24.1 |
| No | 458 | 105 | 22.9 | 22.5 to 23.3 |
| **Self-Reported History of**<br>**previous COVID-19 infection** |  |  |  |  |
| Yes | 88 | 69 | 78.4 | 77.5 to 79.3 |
| No | 600 | 81 | 13.5 | 13.2 to 13.8 |
| Unknown | 11 | 2 | 18.2 | 15.9 to 20.5 |
| **Median Household Income**<br>**(2015) based on FSA** |  |  |  |  |
| \$40-<\$60K | 9 | 1 | 11.1 | 9.1 to 13.2 |
| \$60-<\$80K | 97 | 22 | 22.7 | 21.8 to 23.5 |
| \$80-<\$100K | 570 | 130 | 22.8 | 22.5 to 23.2 |
| \$100K+ | 228 | 56 | 24.6 | 24.0 to 25.1 |
| **Essential workers** |  |  |  |  |
| Yes | 228 | 50 | 21.9 | 21.4 to 22.5 |
| No | 352 | 73 | 20.7 | 20.3 to 21.2 |
| Prefer not to<br>answer | 113 | 21 | 18.6 | 17.9 to 19.3 |
| **Completed Education** |  |  |  |  |
| High school or less | 121 | 31 | 25.6 | 24.8 to 26.4 |
| College, Trade,<br>Certificate | 89 | 18 | 20.2 | 19.4 to 21.1 |
| University degree | 453 | 89 | 19.6 | 19.3 to 20.0 |
| Prefer not to<br>answer | 30 | 6 | 20.0 | 18.6 to 21.4 |
| **Multi-generational**<br>**Household** |  |  |  |  |
| Yes | 125 | 30 | 24.0 | 23.3 to 24.7 |
| No | 451 | 88 | 19.5 | 19.1 to 19.9 |
| Prefer not to<br>answer | 78 | 15 | 19.2 | 18.4 to 20.1 |
| **Location** |  |  |  |  |
| Peel Resident | 830 | 194 | 23.4 | 23.1 to 23.7 |
| Brampton | 742 | 178 | 24.0 | 23.7 to 24.3 |
| Caledon | 39 | 7 | 17.9 | 16.7 to 19.2 |
| Mississauga | 49 | 9 | 18.4 | 17.3 to 19.5 |

\* Test interpretation for vaccinated individuals differed from non-vaccinated individuals, see Methods section Essential worker refers to manufacturing, food processing, transportation sector

**Figure 2:**
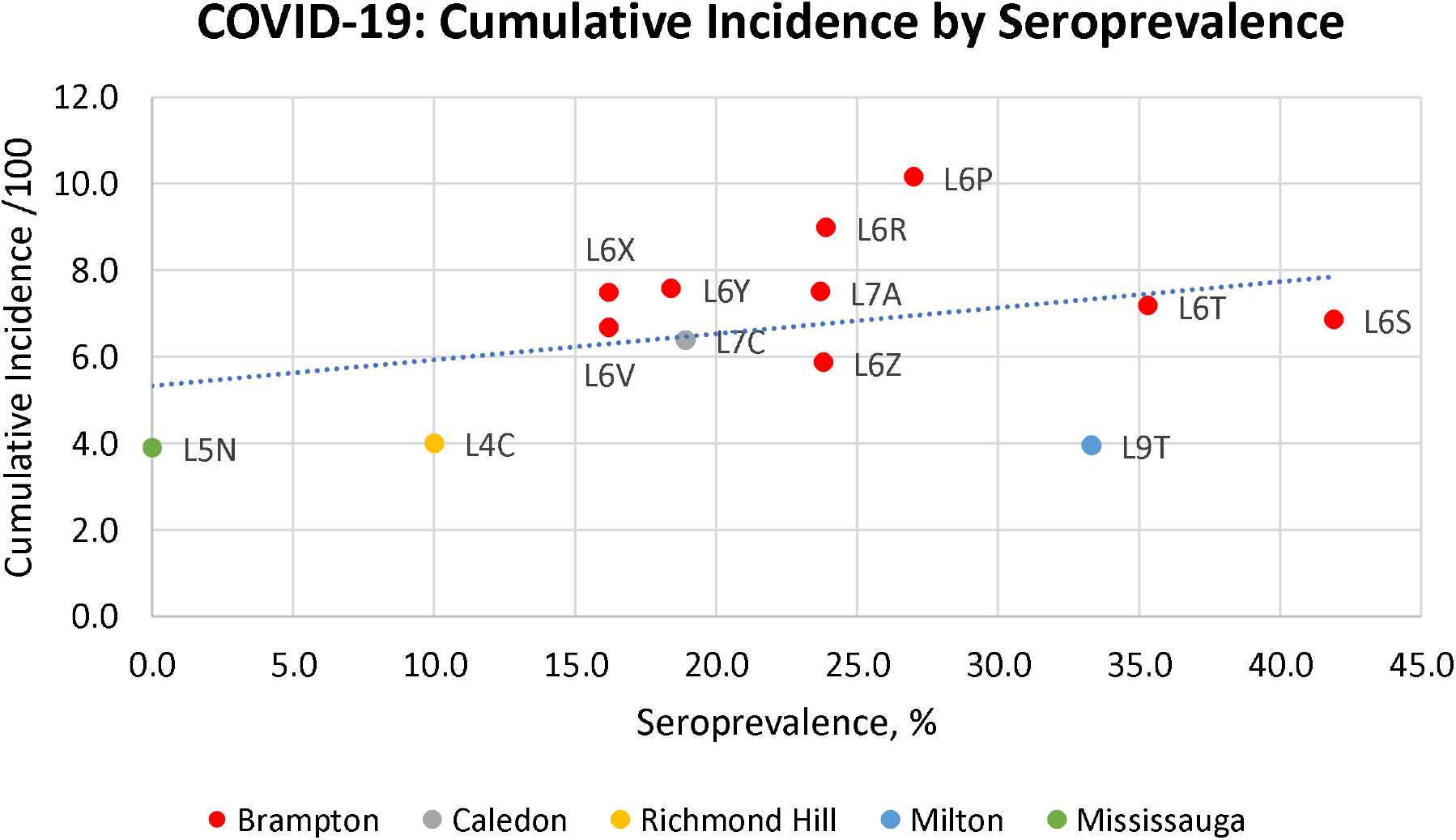
Cumulative Incidence of COVID-19 cases (per 100) as of Oct 3, 2021 by Age-Sex Standardized Seroprevalence by FSA. Cumulative Incidence data from Institute of Clinical Evaluative Sciences (31).

### Risk Perception

Of those who completed the risk perception questionnaire (Table 3), 92% reported COVID-19 posed a major threat to the South Asian community, 51% agreed or strongly agreed that they were at high risk from COVID-19 because of their location, 35% reported being high risk because of their work or profession, 14% because of socializing or lifestyle, and 14% because of their housing situation.

**Table 3:** Risk Perception.

|  | **Proportion**<br>**Agree#** | **Score Mean**<br>**(SD)** |
| --- | --- | --- |
| Number completed secondary survey | 576 |  |
| COVID-19 poses a major threat to our community | 91.5% | 4.5 (0.8) |
| I am at a high risk from COVID-19 because of my location | 51.0% | 3.4 (1.2) |
| I am at a high risk from COVID-19 because of my profession or my work | 35.4% | 2.8 (1.4) |
| If I get exposed to/contract COVID-19, I am likely to have serious symptoms because of age, and/or pre-existing conditions | 20.2% | 2.5 (1.2) |
| The situation around COVID-19 is over exaggerated/overblown | 19.5% | 2.4 (1.2) |
| If I get exposed to/contract COVID-19, I am likely to need hospitalization because of my age, and/or pre-existing conditions | 15.2% | 2.4 (1.1) |
| I am at a high risk from COVID-19 because of my lifestyle (socializing or working in a crowded place) | 14.2% | 2.2 (1.2) |
| I am at high risk from COVID-19 because of my housing situation | 13.9% | 2.2 (1.1) |

Response captured on 5-point scale of strongly disagree=1, disagree, neutral, agree, or strongly agree=5. #Percent refers to combination of Agree (4) and Strongly Degree (5).

### Sources of Health Information

The top ranked health information sources included healthcare providers or provincial public health bodies (52%), traditional media sources (TV news channels, newspapers) (45%), social media (28%), and friends, family or coworkers (26%) (Figure 3).

**Figure 3:**
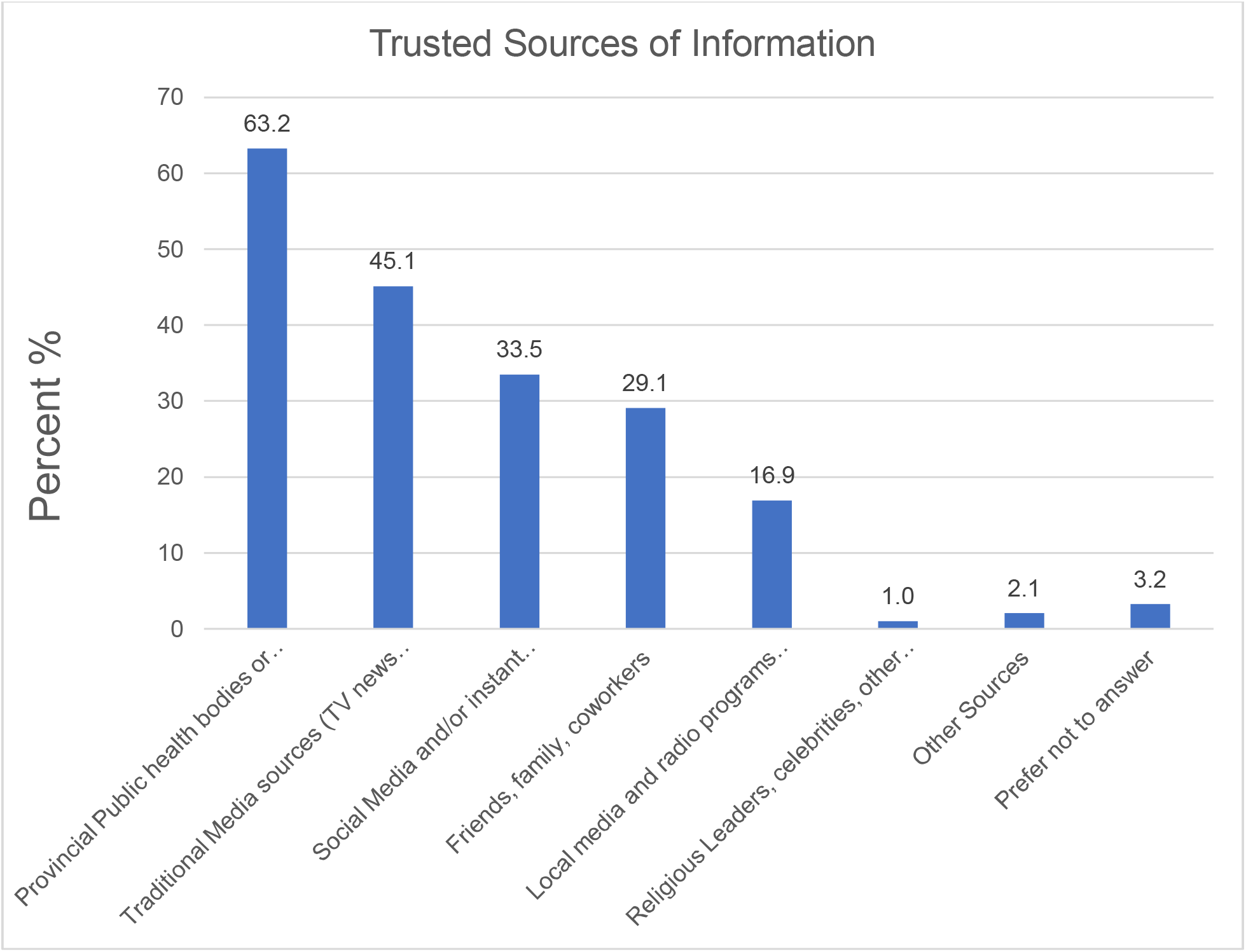
Top 3 most Trusted Sources of COVID Health Information Sources according to respondents (N=585 respondents).

## Discussion

Among a sample of South Asians recruited primarily from a designated hot spot in the GTA, we report a seroprevalence of SARS-CoV-2 of 23.6% (95% CI: 20.8 to 26.4), which is standardized by age and sex to the Ontario South Asian population. The range in seropositivity using the strict definition to lenient definition was 22% to 27%. The risk of bias assessment was classified as moderate, attributed to the non-probability sampling method. This report confirms other sources of evidence that racialized groups in general, as well as South Asians, specifically in the GTA, represent a high-risk group for SARS-CoV-2 infection (7). Since it utilizes serological testing, it captures infection rate more comprehensively than routine data sources, which rely on SARS-CoV-2 infection PCR testing.

Prior to the COVID-19 pandemic, ethnicity data are not routinely collected or made publicly available in Canada (16). As a surrogate, in areas where a single ethnic group is concentrated, infection and hospitalization rates can be examined by postal code, and then inferred to be representative of high-density ethnic populations (6). However, our data provide direct evidence of the high infection rate suffered by South Asians in the Peel Region. The reasons for this are multiple, and include socio-cultural factors including socioeconomic inequity, multigenerational households, barriers to accessing healthcare including SARS-CoV-2 testing, and the higher percentage of workers who were essential workers in factories, manufacturing, Amazon Inc. centres, and the trucking industry (17-19). Immigrants are overrepresented in meat, food and beverage processing plants, trucking, and in health and long-term care as nurses’ aides and orderlies. The reasons for overrepresentation include the low pay and lack of benefits and protections, features not desired by others who can pursue jobs in other sectors (20,21). Almost 33% of participants in our study reported doing essential work, although an additional 12% preferred not to answer this question, and 19% reported living in a multigenerational household, with an additional 12% preferring not to answer. Participants who indicated they “preferred not to answer” may reflect the stigmatization they may have felt during the initial waves of the pandemic. Therefore, the proportion of essential workers and those living in multigenerational households may be underestimates, especially considering our observation that participants who did not complete the follow-up survey where this information was collected, were more likely to be seropositive for SARS-CoV-2 (Supplementary Table 2).

Our seropositivity data mirrors the high infection rate when comparing FSAs in the City of Brampton and Peel Region (Figure 2). The seropositivity rate was higher in men compared to women, those with lower educational attainment, those living in Brampton, and in multigenerational households. Higher seropositivity of 2 to 3-fold has been reported in ethnic minority groups including Black and South Asian people compared to white people living in the United Kingdom (22,23). Compared to population-based seroprevalence studies conducted during the third wave of the pandemic in Canada such as the Canadian Blood Services with a seropositivity for previous infection of 3.95%, we observed a relatively high rate of seropositivity among South Asians in the GTA (23.6%) (24). However, a similarly high rate of seropositivity (22%) was reported among a cross-sectional study of 1,100 incarcerated men in Canada attributed to enhanced daily interactions in a congregate setting (25).

During the initial waves of the pandemic, it was unclear if standard public health messaging by public health officials at the Federal, Provincial, and local levels was reaching communities at high-risk such as South Asians (26). Our data, collected during the third pandemic wave, show that the top sources of information utilized by our cohort included information from healthcare providers or Public Health, traditional media sources, and social media (Figure 3). It is possible that our findings reflect the impact of activities of South Asian advocacy groups to raise awareness of health risks, and testing for SARS-CoV-2 infection, including provision of health information in multiple South Asian languages. For example, at the outset of the second wave of the pandemic, a physician-led group, named the South Asian COVID task force, opened a culturally sensitive testing centre in the Embassy Grand convention centre, with Punjabi signage and information translated into multiple South Asian languages, which was funded for and supported by Peel Public Health (27,28). Another advocacy group, the South Asian Health Network advocated for isolation centres; sick pay and time off work for testing and vaccination for essential workers, and COVID-19 information sessions regularly (29). Finally, Peel Public Health worked closely with community leaders to provide outreach onsite for essential workers, initiate testing at workplaces, and to develop mass vaccine centres with cultural sensitivity (16).

Finally, over 90% of the South Asian participants reported COVID-19 posed a major threat to the South Asian community, almost half reported their risk of acquiring COVID-19 was high due to their location, and more than 40% reported their risk was high due to their type of employment. In comparison, the minority of participants perceived their risk of COVID to be due to their lifestyle, such as socializing, or to their housing situation.

Limitations of our analysis include recruitment from vaccination centres, which may attract more health-conscious individuals, the selection of whom could lead to an underestimate of the true seroprevalence (Supplementary Table 1). Further, the history of COVID-19 infection was based on self-reporting which is less reliable than laboratory confirmed diagnosis.

## Conclusion

The COVID-19 pandemic was particularly devasting for the South Asian community in the GTA, Ontario. In this analysis, we show the seropositivity data confirms a high infection rate of 23.6% from samples collected during the peak of Wave 3. Planning for future COVID-19 waves should incorporate an understanding of socio-cultural determinants of such high-risk communities.

## Data Availability

All data produced in the present study are available upon reasonable request to the authors

## Acknowledgements

We acknowledge the funder of this work COVID-19 Immunity Task Force (CITF), Public Health Agency of Canada. We acknowledge the advocacy support of the South Asian COVID-19 Task Force, and the South Asian Health Network, and Peel Public Health. Dr. Anand holds a Canada Research Chair (Tier 1) Ethnic Diversity and Cardiovascular Disease, and Michael G. DeGroote Heart and Stroke Foundation Chair in Population Health; Dr. Marc-Andre Langlois holds a Canada Research Chair in Molecular Virology and Intrinsic Immunity. Dr. Dawn Bowdish holds a Canada Research Chair in Aging & Immunity.

## Study Team

**Principal Investigator:** Dr Sonia S Anand

**Co-investigators:** Dr. Shrikant Bangdiwala; Dr. Shelly Bolotin; Dr. Dawn Bowdish; Dr. Rahul Chanchlani; Dr. Russell de Souza; Dr. Sujane Kadasamy, Dr. Marc Andre Langlois, Dr. Scott Lear; Dr. Mark Loeb; Dr. Lawrence Loh, Dr. Zubin Punthakee; Dr. Gita Wahi

**Project Office: Program Manager** - Dipika Desai; **Senior Research Coordinator** - Dr. Jodi Miller; **Junior Research Coordinator** - Andrea Rogge, Sherry Zafar; **Statistician** - Karleen Schulze; Database Management: Marsella Bishop, Natalie Williams

**Ontario Recruitment Team:** Site Coordinator: Farah Khan; **Research Assistants** - Zainab Khan, Jayneel Limbachia

**Student Volunteers**: Rabeeyah Ahmed, Archchun Ariyarajah, Riddhi Chabrota, Yajur Iyengar, Lokeesan Kaneshwaran, Elias Larrazabal, Hemantika Mahesh, Baanu Manoharan, Rhea Murti, David Nash, Nisarg Radadia, Khyathi Rao, Anand Sergeant, Rhea Varghese

**Dr. Marc-Andre Langlois Serology Lab**: Corey Arnold, Martin Pelchat, Kiran Nakka.

