## Supplementary Figure 1 for "What factors converged to create a COVID-19 hot-spot? Lessons from the South Asian community in Ontario"

**SFigure 1: Consort Diagram**

**First dried blood spot samples in Ontario**

**N=939**

**Study sample recruited from**

Vaccine Centre (n=848)

Social media, family, friends (n=51)

Vishnu Mandir (n=9)

SAHARA study (n=8)

**Total N=916**

**Exclusions**

DBS not linked to participant (n=12)

Inadequate DBS sample (n=9)

Incomplete key dates (n=2)

**Study sample with completed surveys**

**N=693 (75.7%)**
