## Supplementary Table 1 for "What factors converged to create a COVID-19 hot-spot? Lessons from the South Asian community in Ontario"

**Supplementary Table 1: Risk of bias assessment**

| **Item 1: Was the sample frame appropriate to address the target population?** | | | |
| --- | --- | --- | --- |
| **Yes** | | Sample frame described and it approximated the target population | |
| No | | Sample frame did not approximate the target population (e.g., blood donors do not represent general population, doctors do not represent all health care providers) | |
| Exclude | | Sample frame not described | |
| *Notes | | The term “target population” should not be taken to infer every individual from everywhere or with similar disease or exposure characteristics. Instead, give consideration to specific population characteristics in the study, including age range, gender, morbidities, medications, and other potentially influential factors. For example, a sample frame may not be appropriate to address the target population if a certain group has been used (such as those working for one organisation, or one profession) and the results then inferred to the target population (i.e. working adults). A sample frame may be appropriate when it includes almost all the members of the target population (i.e. a census, or a complete list of participants or complete registry data). | |
| **Item 2: Were study participants recruited in an appropriate way?** | | | |
| Yes | Probability sampling method (simple or stratified random) or entire sample (e.g., an entire town) was used | | |
| **No** | Non-probability sampling | | |
| Exclude | Sampling method not reported | | |
| **Item 3: Was the sample size adequate?** | | | |
| **Yes** | ≥599 | | |
| No | <599 | | |
| Exclude | Sample size not reported | | |
| *Notes | To calculate the required sample size we used an assumed prevalence of 2.5%, which was the global average estimated by the WHO in April, 2020.^1^ Based on guidance by the Joanna Briggs Institute and published medical statistical recommendations we selected a precision value that was half the assumed prevalence (1.25%) [2,3]. We calculated a minimum sample size of 599 using these inputs:  Sample size calculation: $n={\frac{Z^{2}P(1-P)}{d^{2}}}$  Where n = sample size;  Z = Z statistic for level of confidence (95%);  P = expected prevalence (2.5% WHO global estimate);  d = precision (1.25%)  In cases where the sample size calculation was provided and the required sample for 80% power was below our threshold (n<599), this item was marked as yes. | | |
| **Item 4: Were the study subjects and setting described in detail?** | | | |
| **Yes** | Average age and distribution of gender/sex provided | | |
| No | Neither age or gender/sex is provided, or only one of age and gender/sex is provided | | |
| **Item 5: Was data analysis conducted with sufficient coverage of the identified sample?** | | | |
| **Yes** | The demographic characteristics (gender/sex, age, and ethnicity) of the sample is at least somewhat representative of the population | | |
| No | The demographic characteristics (gender/sex, age, and ethnicity) of the sample is not representative of the population | | |
| Unclear | Information is not provided about demographic characteristics of the sample (gender/sex, age, and ethnicity) | | |
| **Item 6: Were valid methods used for the identification of the condition?** | | | |
| **Yes** | The test used met the FDA standards for Emergency Use Authorizations for COVID-19 serological tests: sensitivity minimum 90%, specificity minimum 95%, as reported in the study [4]. | | |
| No | The test used did not meet the FDA standards for Emergency Use Authorizations for COVID-19 serological tests: sensitivity minimum 90%, specificity minimum 95%. | | |
| Exclude | Test sensitivity and specificity not reported | | |
| **Item 7: Was the condition measured in a standard, reliable way for all participants?** | | | |
| **Yes** | The same serology test was used for all participants | | |
| No | Different serology tests were used for participants | | |
| Unclear | No details were provided about which participants received which serology tests | | |
| **Item 8: Was there appropriate statistical analysis?** | | | |
| **Yes** | Does all of the following: corrects for population characteristics or the sample is somewhat representative of the population (probability sampling), corrects for test characteristics), and provides the information necessary to determine the numerator, denominator, prevalence estimate, and confidence interval. | | |
| No | Does not correct for population characteristics and the sample is not likely representative of the population (non-probability sampling), does not correct for test or provide the information necessary to correct for test characteristics, or does not provide the information necessary to determine the numerator, denominator, prevalence estimate, and confidence interval. | | |
| **Item 9: Was the response rate adequate, and if not, was the low response rate managed appropriately?** | | | |
| **Yes** | Response rate > 60% or the demographics of the sample were a reasonable match to those of the target population [5] | | |
| No | Response rate < 60% and the demographics of the sample were not a reasonable match to those of the target population | | |
| Unclear | Response rate not provided and it was unclear if the demographics of the sample differed from the target population | | |
| **Item 10: Overall risk of bias** | | | |
| Low | | | The estimates are very likely correct for the target population. To obtain a low risk of bias classification, all criteria must be met or departures from the criteria must be minimal and unlikely to impact on the validity and reliability of the prevalence estimate. These include sample sizes that are just below the threshold when all other criteria are met, reporting only some of characteristics of the sample, test characteristics below the threshold but corrections for the test performance, and response rates that are just below the threshold in the context of probability-based sampling of an appropriate sampling frame with population weighted seroprevalence estimates. |
| **Moderate** | | | The estimates are likely correct for the target population. To obtain a moderate risk of bias classification, most criteria must be met and departures from the criteria are likely to have only a small impact on the validity and reliability of the prevalence estimates. |
| High | | | The estimates are not likely correct for the target population. To obtain a high risk of bias, many criteria must not be met or departures from criteria are likely to have a major impact on the validity and reliability of the prevalence estimates. |
| Unclear | | | There was insufficient information to assess the risk of bias. |
