## Supplementary Table 2 for "What factors converged to create a COVID-19 hot-spot? Lessons from the South Asian community in Ontario"

**Supplementary 2: Demographics in Responders versus Non-responders**

|  | | **Completed Survey** | |
| --- | --- | --- | --- |
|  | **Overall** | **Yes** | **No** |
| **N** |  | 693 | 223 |
| Female | 49.2% | 50.2% | 46.2% |
| Age | 41.5 | 40.1 | 45.7 |
| FSA median household income >=$80,000 | 88.3% | 88.5% | 87.5% |
| Brampton Resident | 82.0% | 81.6% | 83.3% |
| Peel Resident | 91.7% | 91.7% | 91.7% |
| Seroprevalence | 23.1% | 20.8% | 30.5% |

Note: Those that did not complete the survey were slightly older, while slightly more females completed.

Among the non-completers, the seroprevalence was 10% higher.
