## Supplementary Table 3 for "What factors converged to create a COVID-19 hot-spot? Lessons from the South Asian community in Ontario"

**Supplementary Table 3: Demographics by Vaccination^#^ status**

|  | **Pre-vaccination Group** | **Vaccinated Group** | **Responses** |
| --- | --- | --- | --- |
| **Overall** | 458 | 458 | 916 |
| **Sex** |  |  | 916 |
| Female | 234 (51.1) | 217 (47.4) |  |
| Male | 223 (48.7) | 239 (52.2) |  |
| Self-described | 1 (0.2) | 2 (0.4) |  |
| **Age group** |  |  | 906 |
| 18-24 | 54 (11.9) | 41 (9.1) |  |
| 25-34 | 162 (35.7) | 71 (15.7) |  |
| 35-44 | 154 (33.9) | 93 (20.6) |  |
| 45-54 | 55 (12.1) | 102 (22.6) |  |
| 55-64 | 22 (4.8) | 72 (15.9) |  |
| 65+ | 7 (1.5) | 73 (16.2) |  |
| **Vaccinated*** |  |  | 916 |
| No | 458 (100.0) | 0 (0.0) |  |
| Yes | 0 (0.0) | 458 (100.0) |  |
| 1 dose | 0 (0.0) | 393 (85.8) |  |
| 2 doses | 0 (0.0) | 65 (14.2) |  |
| **History of previous COVID-19 infection** |  |  | 699 |
| Yes | 47 (13.2) | 41 (12.0) |  |
| No | 299 (84.0) | 301 (87.8) |  |
| Unknown | 10 (2.8) | 1 (0.3) |  |
| **Median Household Income (2015) based on FSA** |  |  | 904 |
| $40-<$60K | 3 (0.7) | 6 (1.3) |  |
| $60-<$80K | 44 (9.7) | 53 (11.8) |  |
| $80-<$100K | 311 (68.7) | 259 (57.4) |  |
| $100K+ | 95 (21.0) | 133 (29.5) |  |
| Prefer not to answer | 0 (0.0) | 0 (0.0) |  |
| **Essential workers** |  |  | 693 |
| Yes | 101 (27.2) | 127 (39.4) |  |
| No | 194 (52.3) | 158 (49.1) |  |
| Prefer not to answer | 76 (20.5) | 37 (11.5) |  |
| **Completed Education** |  |  | 693 |
| High school or less | 68 (18.3) | 53 (16.5) |  |
| College, Trade, Certificate | 43 (11.6) | 46 (14.3) |  |
| University degree | 237 (63.9) | 216 (67.1) |  |
| Prefer not to answer | 23 (6.2) | 7 (2.2) |  |
| **Multi-generational Household** |  |  | 654 |
| Yes | 65 (18.7) | 60 (19.6) |  |
| No | 234 (67.2) | 217 (70.9) |  |
| Prefer not to answer | 49 (14.1) | 29 (9.5) |  |
| **Years in Canada** |  |  | 717 |
| 10 years or less | 142 (39.4) | 81 (22.7) |  |
| >10 years | 155 (43.1) | 229 (64.1) |  |
| Born in Canada | 44 (12.2) | 41 (11.5) |  |
| Prefer not to answer | 19 (5.3) | 6 (1.7) |  |
| **Mother tongue*** |  |  | 732 |
| Punjabi or Urdu | 190 (52.5) | 173 (46.8) |  |
| Hindi | 37 (10.2) | 46 (12.4) |  |
| Gujarati | 44 (12.2) | 62 (16.8) |  |
| Other South Asian Languages | 85 (23.5) | 67 (18.1) |  |
| English | 14 (3.9) | 40 (10.8) |  |
| Prefer not to answer | 4 (1.1) | 0 (0.0) |  |
| **Medical history** |  |  | 679 |
| CVD (MI/Angioplasty/Stroke) | 1 (0.3) | 20 (6.3) |  |
| **Chronic Medical condition requiring medication** |  |  | 666 |
| Hypertension | 14 (4.0) | 34 (10.9) |  |
| Diabetes | 14 (4.0) | 39 (12.5) |  |
| Arthritis | 6 (1.7) | 4 (1.3) |  |
| Chronic Lung disease | 0 (0.0) | 1 (0.3) |  |
| Cancer | 0 (0.0) | 1 (0.3) |  |
| **Smoking Status** |  |  | 641 |
| Never | 294 (88.0) | 261 (85.0) |  |
| Former | 24 (7.2) | 29 (9.4) |  |
| Current | 16 (4.8) | 17 (5.5) |  |
| **Location** |  |  | 905 |
| Region of Peel | 441 (97.4) | 389 (86.1) |  |
| City of Brampton | 410 (90.5) | 332 (73.5) |  |
| City of Caledon | 21 (4.6) | 18 (4.0) |  |
| City of Mississauga | 10 (2.2) | 39 (8.6) |  |

Presented data are n (%). *Multiple answers can be selected.

CVD: Cardiovascular Disease

#Vaccinated group includes 393 (85.8%) with a single dose and 65 (14.2%) with two vaccine doses.
